# Reconstructing the silent circulation of West Nile Virus in a Caribbean island during 15 years using sentinel serological data

**DOI:** 10.1101/2025.02.07.25321837

**Authors:** Celia Hamouche, Jennifer Pradel, Nonito Pagès, Véronique Chevalier, Sylvie Lecollinet, Jonathan Bastard, Benoit Durand

**Affiliations:** EPIMIM, Laboratoire de Santé Animale, ANSES, Ecole Nationale Vétérinaire d’Alfort, 94700 Maisons-Alfort, France; UMR ASTRE, CIRAD, INRAe, Université de Montpellier, Montpellier, France; ASTRE, CIRAD, Petit-Bourg, Guadeloupe; ASTRE, CIRAD, Antananarivo, Madagascar; Sorbonne Université, INSERM, IPLESP, Paris, France

**Keywords:** West Nile, model, vector-borne, sentinel, surveillance, serology, Caribbean, Bayesian

## Abstract

The dynamics of zoonotic infectious diseases with silent circulation may be imperfectly understood and monitored using passive (or reactive) epidemiological surveillance data only, highlighting the interest of quantitative methods like modelling. West Nile virus (WNV) is a widespread mosquito-borne virus transmitted from birds to “dead-end” hosts including humans and horses, in whom it can be fatal. It was first detected in Guadeloupe archipelago, Caribbean, in 2002, although no WNV clinical case in humans nor horses had been reported before 2024. Undetected infections represent a risk as WNV can be transmitted *via* blood and organ donations. In Guadeloupe, epidemiological surveillance started in 2002 in sentinel chickens and horses and in 2015 in mosquitoes, to detect WNV and to improve knowledge on its epidemiology and dynamics. In order to reconstruct the WNV force of infection (FOI), we built a model assessing different hypotheses regarding its dynamics using serological results in respectively 1,022 and 3,649 blood samples collected from 256 horses and 317 chickens between 2002 and 2018. We fitted the model to the serological data using Markov Chains Monte Carlo. We found that WNV FOI in Guadeloupe Island presented both within-year (seasonal) and between-years fluctuations. We identified three main episodes of WNV circulation on the island between 2002 and 2017. During years with circulation, the FOI was predicted to be highest around the months of October-November, although transmission could occur all year long. We estimated a very low weekly seroreversion rate, which is consistent with a lifelong persistence of WNV IgG antibodies in many infected individuals. To conclude, combining longitudinal serological data to a mathematical model allowed reconstructing the recurrent and silent circulation of WNV in this Caribbean island, which could improve surveillance design for better virus detection.

## Introduction

Sentinel (active or proactive) surveillance is defined as the repeated collection of information from same selected individuals or groups to identify changes in the health status of a specified population over time [1]. It is complementary to passive (reactive or clinical) surveillance designs where health adverse events are reported by stakeholders (e.g. hospitals, veterinarians, …) as part of their usual activities [2–4]. Although it often requires substantial resources, active surveillance has the advantage to provide a less biased and more complete picture of an infection occurrence [3,5]. It is particularly useful for pathogens that are under-reported by passive surveillance, for instance when asymptomatic infections are frequent as with arboviruses [6,7] or in settings with limited routine surveillance capabilities [8]. Active surveillance is also adapted to zoonotic diseases arising from wildlife, because human infections then result from incidental transmissions from an animal reservoir source with generally less known demographic (movements, interactions between individuals and populations) and epidemiological (pathogen prevalence and mortality) patterns [9]. When such wildlife zoonotic pathogens circulate endemically, it then becomes appropriate to monitor infections in sentinels from better-followed populations such as domestic animals [10–12].

West Nile virus (WNV), an *Orthoflavivirus* transmitted by mosquitoes mostly of the *Culex* genus, meets most of these criteria. Indeed, wild birds are primary WNV reservoirs, although the virus can spread to mammals including horses and humans [13]. In both species, WNV infection is most often asymptomatic but may result in febrile forms (dengue-like symptoms in humans) and, in some cases, in severe neurological symptoms sometimes leading to death. These species are considered “dead-end hosts” since biting mosquitoes cannot get infected after feeding on them nor further transmit the virus [13,14]. However, WNV can still spread among humans through blood transfusions and organ transplantations from asymptomatic infected donors [15,16]. It is therefore of interest for both human and animal health to monitor its circulation over time and space. This is why simultaneous multi-host surveillance of this pathogen has been emphasized [6,17]. Indeed, WNV sentinel surveillance in many countries has been implemented in multiple host species such as horses, wild and domestic birds or zoo animals, and in vectors [18–34].

In the Americas, WNV was first reported in New York (United States) in 1999, and subsequently spread to the rest of North America, Latin America and the Caribbean [35,36]. Guadeloupe archipelago (French West Indies, Caribbean) has a tropical climate and is populated by ~384,000 inhabitants. WNV circulation in this island was first documented in 2002 when anti-WNV antibodies were found in horses [37]. Following this discovery, a surveillance program was implemented in humans, horses, chickens and mosquitoes using several designs, namely serosurveys, active, sentinel (including based on risk areas) and passive surveillance [38]. Although no clinical case in humans nor horses was reported on the island until 2024 [39,40], anti-WNV antibodies were occasionally detected in horses and chickens throughout two decades [38,41], suggesting its silent circulation.

Mathematical and statistical models may allow inferring the dynamics of pathogens’ circulation using serological data as markers of past infection in hosts [42–44]. Such models were used to infer on transmission patterns of other mosquito-borne viruses, such as Zika, Japanese Encephalitis, Dengue or Chikungunya viruses [45–51]. Previous studies also fitted or validated mechanistic models of WNV transmission to serological data [52–54], although not using more than two years of data.

Here, our objective was to quantify the level of silent circulation of WNV on Guadeloupe Island between 2002 and 2017. We developed a Bayesian model fitted to longitudinal serological data collected in sentinel chickens and horses, to reconstruct both within-year (seasonal) and between-years variations in the WNV force of infection and to estimate key parameters of its epidemiology and diagnostic.

## Materials and Methods

### Serological surveillance data

For this study, we analyzed longitudinal serological data collected as part of a sentinel surveillance scheme in domestic animals. WNV serological statuses were determined from 1,022 sera sampled from 256 horses in 11 riding clubs between July 2002 and February 2018, and from 3,649 sera sampled from 317 chickens in five chicken farms between November 2013 and August 2018 (Figure 1).

**Figure 1.**
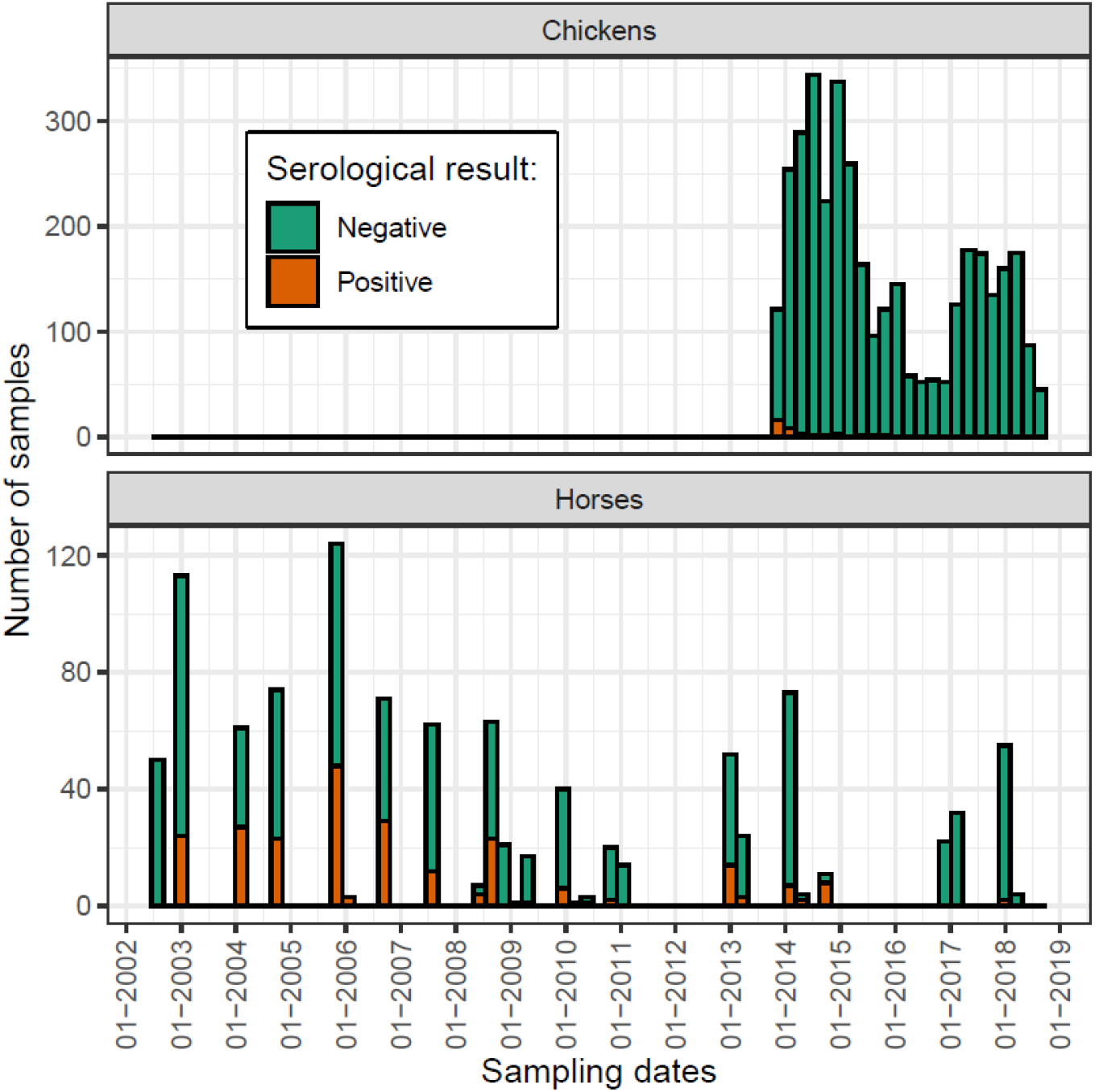
Longitudinal serological data (anti-WNV IgG antibodies) collected from horses and chickens in Guadeloupe between July 2002 and August 2018.

Anti-WNV IgG antibodies were detected in sera using inhibition or competition enzyme-linked immunosorbent assays (Epitope Blocking ELISA, targeting either anti-NS1 or E antibodies) as previously described [37,55,56]. Threshold values defining ELISA-positives were as specified by the manufacturer for the ELISA E commercial kit, or as determined during the development and validation for the ELISA NS1 [57]. Positive samples were then tested by virus neutralization test at the French Reference Laboratory (ANSES).

We did not have the information on animals’ age at sampling to compute their past exposure to the virus. Therefore we used the time between consecutive samples taken from same individuals (i.e. pairs of samples) to infer the virus’ force of infection (FOI) most likely to explain serological transitions (e.g. seroconversions). The median duration between consecutive samples in same individuals was 376 days, i.e. 54 weeks, in horses (interquartile range [306; 593]) and 14 days in chickens (IQR [14; 14]). We discarded two pairs of samples separated by more than 5 years, because we could not reasonably exclude that these animals had undergone more than one serological transition during that period (seroconversion followed by a seroreversion, or the opposite).

### Entomological surveillance data

An entomological surveillance program was set up bi-monthly from November 2015 using CDC CO_2_ mosquito traps (John W. Hock Company, Gainesville, FL) at four sites located near sentinel chicken farms to monitor mosquito populations abundance [38,58]. The entomological data used in this study was the abundance of *Culex* mosquitoes. To better capture the time dynamics of vector populations, we used all data available even beyond the period studied – hence collected between November 2015 and May 2021.

### Ethics statement

Animal samplings have been performed following guidelines and legislation applicable to the surveillance of animal and public health risks (Regulation (EU) 2016/429 of the European Parliament and of the Council of 9 March 2016 on transmissible animal diseases and amending and repealing certain acts in the area of animal health (‘Animal Health Law’)); they have been performed by veterinarians with sanitary authorizations upon request of the veterinary services (DAAF971).

### Serological model

Our model aimed to predict the true WNV serological status *S*_*i,k*_ (valued 0 and 1 for negative and positive status, respectively) of the k^th^ sample taken from individual *i*. For any *k* ≥ 2, *S*_*i,k*_ followed a Bernoulli drawing of probability *p*_*i,k*_:

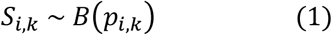

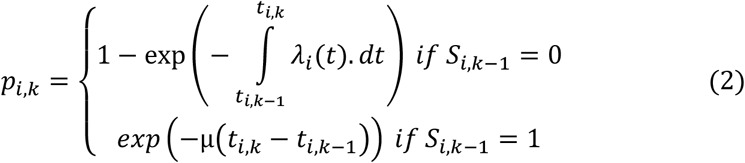

Where *t*_*i,k*_ was the week when sample *k* in individual *i* was collected, *λ*_*i*_*(t)* was WNV FOI that applied to individual *i* on the week *t*, and *μ* was the seroreversion rate. We assumed that *μ* was constant over time and had the same value for horses and chickens. Moreover, we supposed that the FOI could vary within each year between a baseline and a maximum value (peak height). The maximum FOI was assumed to occur on the same week every year but its value depended on the year. The seasonal variations of *λ*_*i*_ were represented by a sinusoid expressed as:

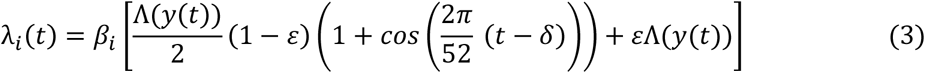

Where β_i_ was the relative risk of WNV infection in an individual *i* compared to a horse individual, with β_i_ = 1 if individual *i* was a horse, and β_i_ = β if it was a chicken. *y(t)* corresponded to the year of week *t* and *Λ(y(t))* was the maximum FOI reached on year *y(t). ε* was the fraction of the FOI that did not vary over the year, hence *εΛ(y(t))* was the baseline FOI reached on year *y(t). δ* was the week of the year when the peak of FOI was reached (Table 1).

**Table 1.**
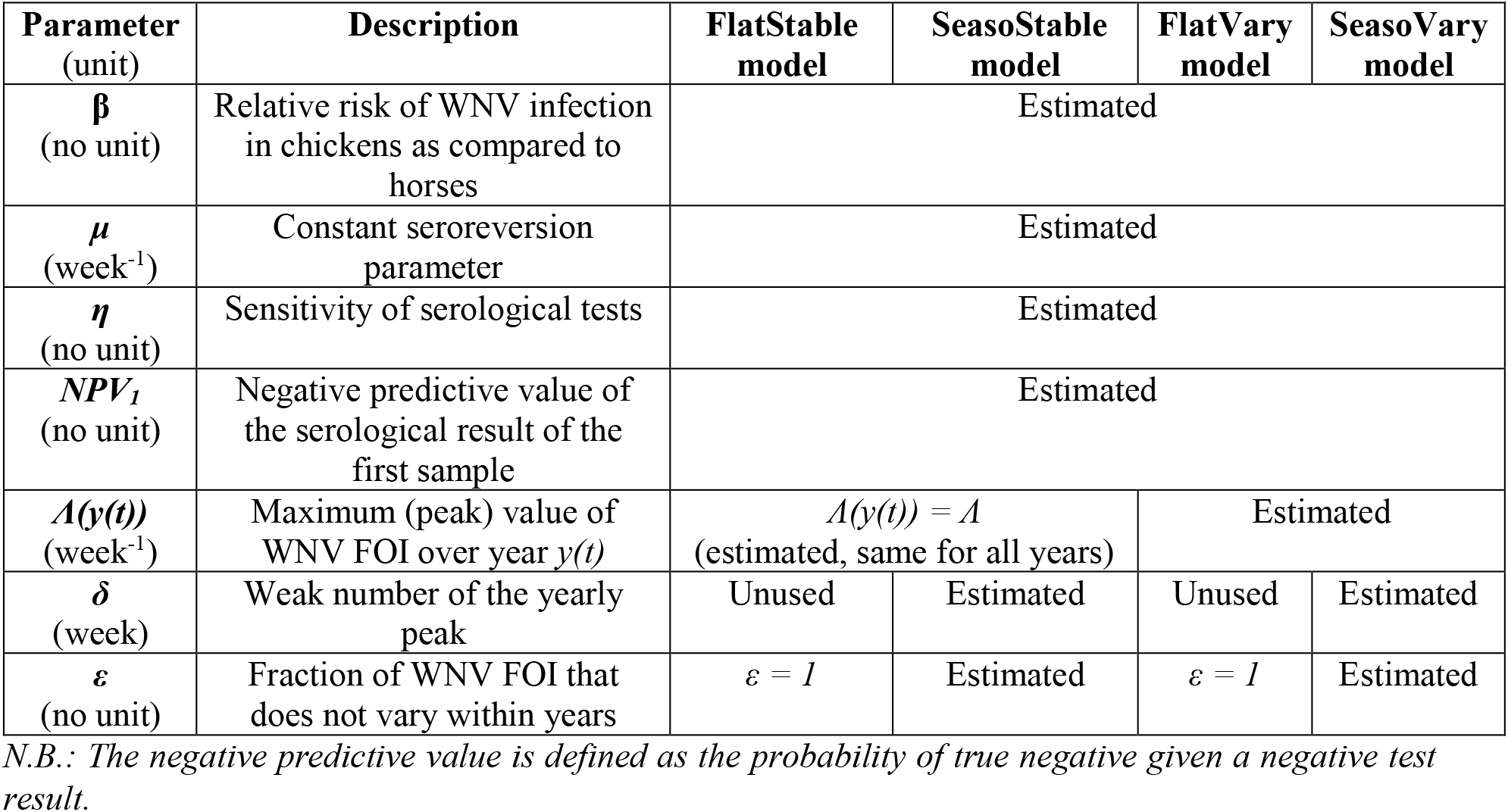
Description of the serological and observation model parameters in the four scenarios. Some parameters were only used in some of the model scenarios.

We defined four scenarios for the model, depending on whether the FOI varied over time, within and/or between years (Figure 2 and Table 1). In “FlatStable” and “FlatVary” models, *ε* was forced to 1, meaning that the seasonal (i.e. within-year) variations of the FOI were ignored. In “SeasoStable” and “SeasoVary” models, ε was estimated. In “FlatStable” and “SeasoStable” models, *Λ(y(t)) = Λ* for all weeks *t*, meaning that we ignored between-year variations. Formulae for each model scenario are summarized in Supplementary Table S1.

**Figure 2.**
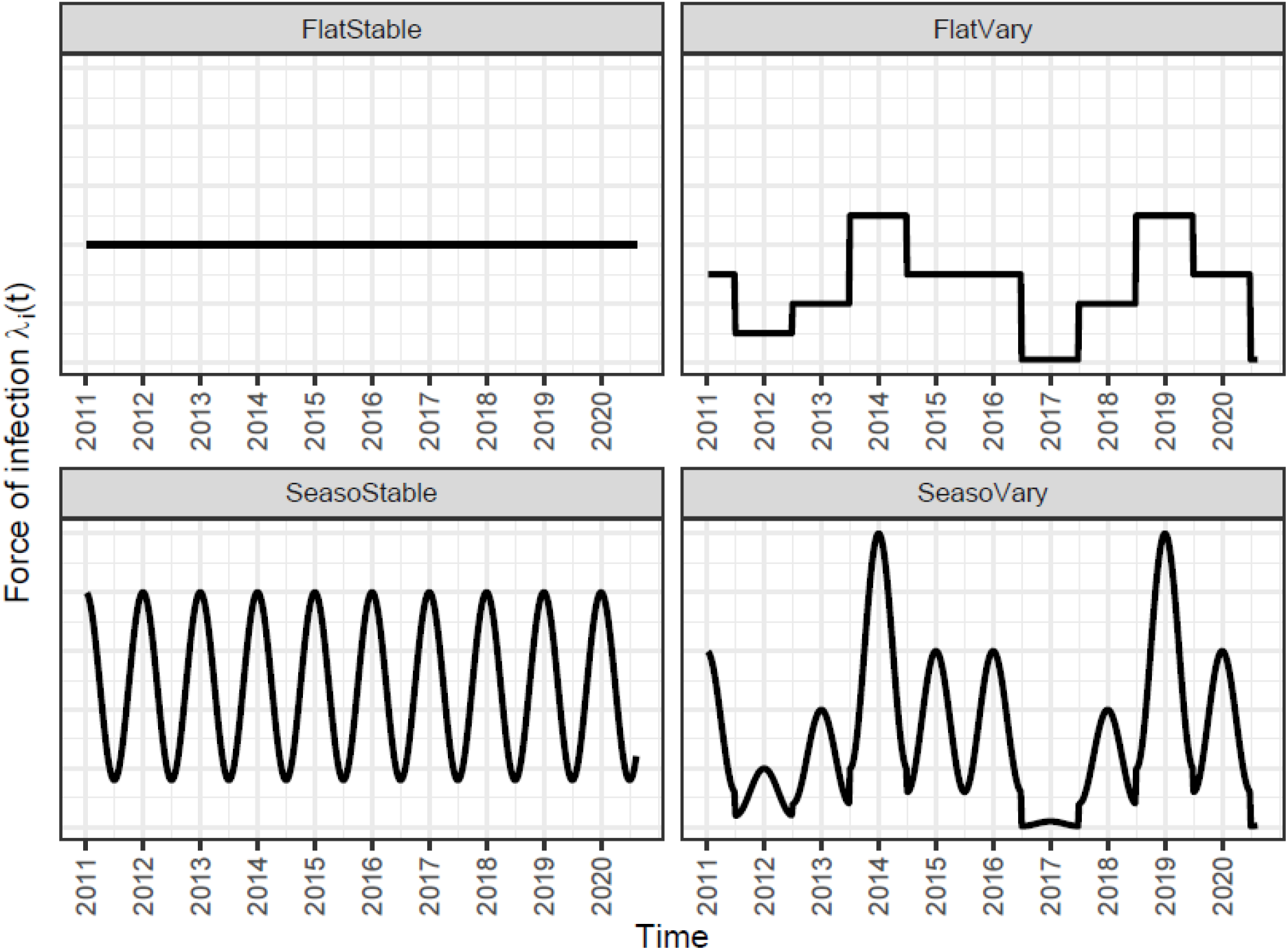
Illustration (synthetic data) of the serological model scenarios used in the study. “SeasoStable” and “FlatVary” models accounted respectively for only within-year (seasonal) and only between-years variations of the force of infection (FOI). “SeasoVary” model accounted for both within- and between-years variations of the FOI. “FlatStable” model did not account for any variation of FOI with time.

### Observation model

The serological result *Y*_*i,k*_ (observed WNV serological status) depended on the test sensitivity *η*, which we estimated, and on the specificity, which we assumed to be 1 (Table 1). Indeed, virus neutralization test is considered the gold standard serological test regarding specificity [56,59]. For any sample *k* ≥ 2 in any individual i:

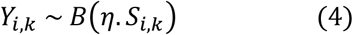

We did not have the information on animals’ age – hence on their previous exposure to the virus – when they were first sampled. Therefore, in order to initialize the model and infer the true serological status of the first sample in each individual *i*, we introduced a parameter *NPV*_*1*_, the negative predictive value of the first sample result for any individual, i.e. the probability of true negative given a first negative test result (Table 1). It is formally defined as *P(S*_*i,1*_*=0*|*Y*_*i,1*_*=0). NPV*_*1*_ depended on *η* and varied according to hyperparameters, as detailed in the Supplementary Note 1. Moreover, since the specificity was assumed to be 1, the positive predictive value (probability of true positive given a positive test) was also 1. Therefore, for any individual *i*:

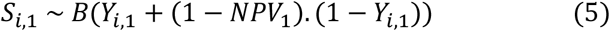

### Models fitting and selection

Model parameters were estimated using a Markov Chain Monte Carlo (MCMC) algorithm, implemented with the R package *rjags* [60]. In this Bayesian framework, most prior distributions were uninformative, although not for parameters *ε* and *δ*. Indeed, we assumed that seasonal variations in WNV FOI (determined by *ε* and *δ*) are partly related to seasonal variations in mosquito abundance. Therefore, we performed the model’s fitting in two steps. In Step 1, we fitted a model analogous to the “SeasoStable” model to the weekly mosquito abundance data, which allowed estimating posterior distributions for seasonality parameters *ε* and *δ* (see details in the Supplementary Note 2). These two distributions were then used as informative priors in Step 2, where the four models were fitted to the serological data (Supplementary Table S2 and Supplementary Figure S1). The usual MCMC convergence diagnostics were performed in both steps. Serological model scenarios were then compared using the Deviance Information Criterion (DIC) and the best fitting model was selected based on the smallest DIC [61].

## Results

### Surveillance results

We analyzed the presence of anti-WNV IgG antibodies in domestic animals sampled as part of a sentinel surveillance scheme in Guadeloupe. Among 764 consecutive pairs of samples collected from horses between 2002 and 2018, 82 (10.7%) seroconversions and 9 (1.2%) seroreversions were observed (Table 2). Among 3,332 consecutive pairs of samples collected from chickens between 2013 and 2018, 6 (0.2%) seroconversions and 6 (0.2%) seroreversions were observed (Table 2).

**Table 2.**
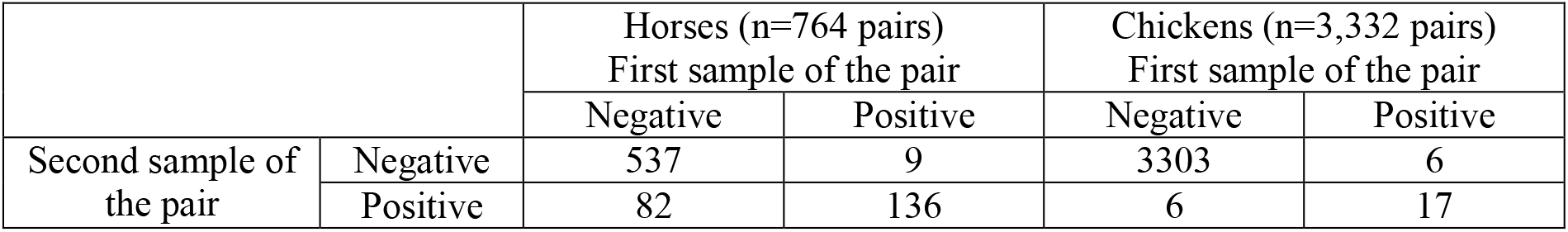
Observed serological results (anti-WNV IgG antibodies) in consecutive pairs of samples collected from horses and chickens in Guadeloupe Island between 2002 and 2018.

*Culex* mosquito abundance data collected bi-monthly between 2015 and 2021 is displayed in Supplementary Figure S2. It showed seasonal patterns that were used to derive informative priors for fitting the serological model.

### Model predictions

After fitting the seasonal model to the mosquito abundance data (Step 1), we fitted the four serological models to the longitudinal serological data (Step 2). The model scenario with the lowest DIC was “SeasoVary” (Supplementary Table S3), suggesting both within- and between-years variations of WNV FOI in Guadeloupe Island. This model predicted that three main episodes of WNV circulation occurred on the island between 2002 and 2017 (see Figure 3): an important one in 2002, followed by another one of smaller intensity in 2007, and finally in 2010-2012, although uncertainty in outbreak intensity (amplitude) was greater for the latter due to less serological data collected.

**Figure 3.**
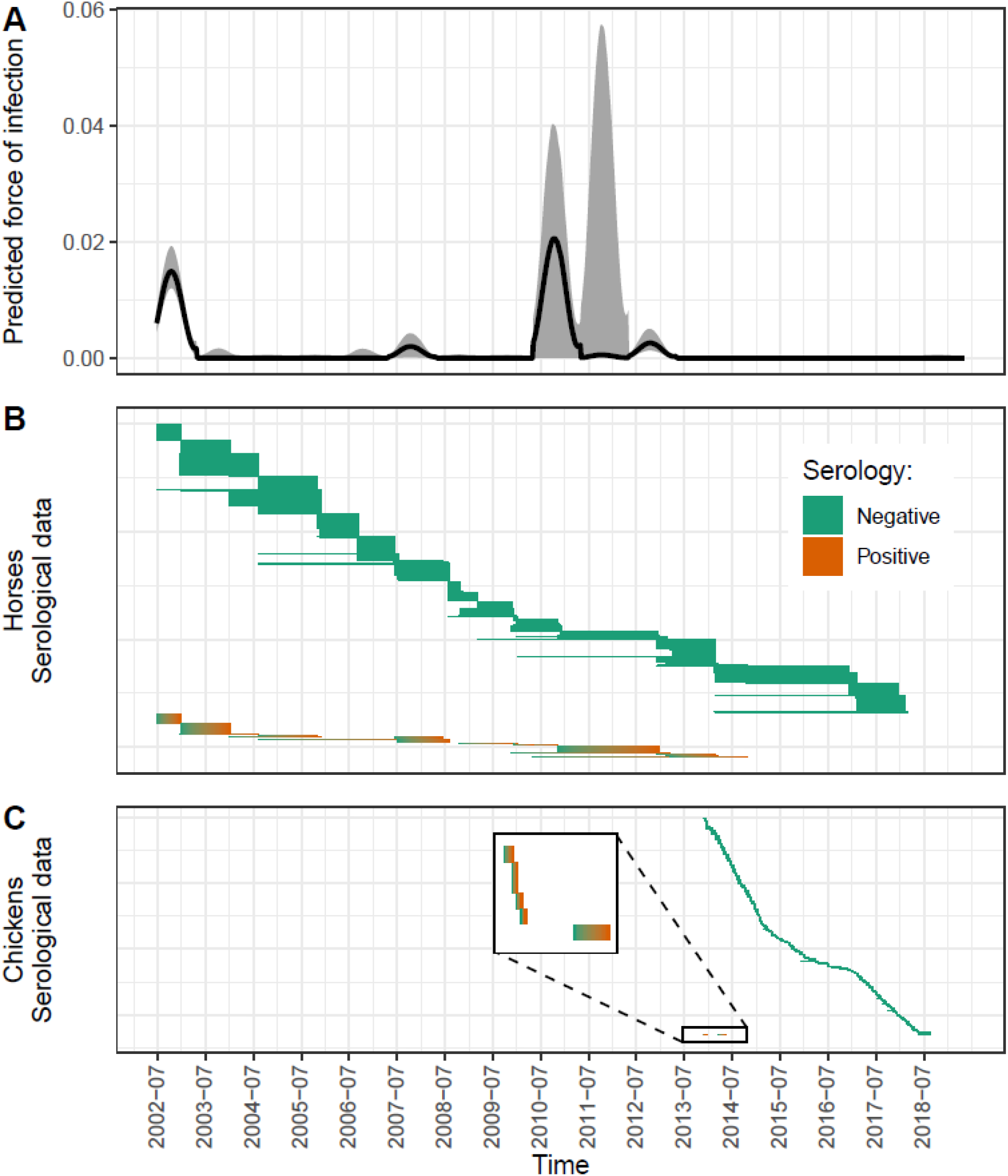
West Nile virus force of infection (FOI) in Guadeloupe Island predicted between 2002 and 2017 by the “SeasoVary” serological model (panel A), and longitudinal serological data collected in horses (panel B) and chickens (panel C). In panel A, the black line represents the median of predictions (using 5,000 repetitions of the model), while the gray area represents the 80% prediction interval. In panels B and C, each row is a pair of consecutive blood samples, and only the observed Negative-Positive and Negative-Negative serological transitions are displayed.

### Parameter estimates

Following Step 1 (fit of the seasonal model to the mosquito abundance data), the median of *δ* was estimated to week 45.3 (95% credible interval: [43.5; 47.4]), implying that the yearly peak of *Culex* mosquito abundance in Guadeloupe Island occurs around October-November (Supplementary Figure S2). Then, after Step 2 (fit to serological data) and for the “SeasoVary” model, the median posterior estimate of *δ* was slightly earlier (week 44.8 [43.0; 46.8]), suggesting a marginal time shift for the yearly peak of WNV FOI. Nevertheless, *ε* was estimated to 0.102 [0.019; 0.226], showing the potential of WNV to circulate all year long in Guadeloupe (Table 3 and Supplementary Figure S3).

**Table 3.** Posterior estimates of parameters of the “SeasoVary” serological model: median and 95% highest posterior density interval (HPDI, credible interval). Posterior distributions for Λ(y(t)) are depicted in Supplementary Figure S3.

| Parameter | Unit | Median of the posterior and 95% credible interval (HPDI) |
| --- | --- | --- |
| $\beta$ | - | 2.50 [0.008 ; 9.24] |
| $\varepsilon$ | - | 0.102 [0.019 ; 0.226] |
| $\delta$ | week | 44.8 [43.0 ; 46.8] |
| $\mu$ | week<sup>-1</sup> | $1.65 \times 10^{-3}$ [$4.37 \times 10^{-4}$ ; $2.87 \times 10^{-3}$] |
| $\eta$ | - | 0.803 [0.676 ; 0.912] |
| $NPV_I$ | - | 0.996 [0.994 ; 0.998] |

We were not able to estimate β, the relative risk of infection in chickens as compared to horses, since its posterior distribution (median of 2.50 [0.008; 9.24]) was almost similar to its prior distribution (Supplementary Figure S3), reflecting a lack of information in the data regarding this parameter. We estimated the seroreversion parameter to 1.65×10^−3^ [4.37×10^−4^; 2.87×10^−3^] per week, i.e. 1/607 weeks or 1/11.7 years.

Furthermore, we estimated the sensitivity *η* to be 0.803 [0.676; 0.912] and *NPV*_*1*_ to be 0.996 [0.994; 0.998], suggesting a good reliability of serological tests (Table 3 and Supplementary Figure S3). This is comparable to values found by [56,62,63].

## Discussion

Some infectious diseases that are transmitted to humans from wildlife reservoir sources, such as WNV, may not be well detected by passive surveillance systems, especially when asymptomatic forms are frequent, sparking a possible silent circulation of the pathogen. In this study, we quantified this silent circulation for WNV in Guadeloupe Island using a serological model fitted to sentinel surveillance data collected in horses and chickens. Among different hypothesis on the variations of WNV force of infection (FOI), the best selected model was “SeasoVary”, suggesting that this FOI changes both within-year and between-years on the island.

Several ecological and epidemiological mechanisms could explain between-years FOI variations. First, variations of climatic factors across years – including occurrence of extreme weather events – may cause heterogeneity in vectors abundance, species composition and/or competence resulting in dramatic changes of epidemiological patterns [54,64–68]. Second, wild bird population renewal over the years may lead to decreasing levels of herd immunity, hence allowing WNV outbreaks to occur again every few years [69]. Third, WNV infection prevalence in migratory birds might vary across years, leading to hypothetical between-years fluctuations in the virus’ introduction risk [70].

Moreover, within-year variations in FOI might also be attributed to various factors. First, we found that the abundance of *Culex* spp. mosquitoes – main WNV vectors – can vary seasonally in such a tropical climate, which was expected [71,72]. Additionally, the distribution of species within the *Culex* genus (including potential enzootic and bridge vectors which are yet to be fully characterized in Guadeloupe) may also change seasonally and may have implications on WNV transmission [73,74]. Second, mechanisms other than mosquito abundance might affect the timing of WNV FOI peak over the year. Indeed, the FOI is also driven by the infection prevalence in vectors, which depends on the seasonal dynamics of infection in wild birds, among other factors. They may depend on demographic traits such as the hatching season, which abruptly supplements the population in susceptible individuals [75]. Furthermore, bird populations on the island fluctuate according to migrations that are highly seasonal, with for instance shorebirds species flying from North America to Guadeloupe Island around August-October [76]. Migratory birds might either seasonally introduce the virus, and/or change the total density of susceptible individuals in the island’s wild bird population [52,77–79]. Seasonal patterns were also observed from sentinel chickens in Florida (United States), where seroconversions mostly occurred between July and August [23].

A limitation of this work is that we did not account for spatial heterogeneity of WNV FOI on the island. However, given Guadeloupe surface area (1,628 square kilometers) and the time between successive blood samples in horses (median of 54 weeks), we cannot preclude that individuals were exposed to mosquito bites in unrecorded locations other than their stable, as part of horse riding tours, competitions or other events related to the equine industry [80].

Previous studies showed the frequent persistence of neutralizing antibodies for at least several months or years in birds [81–84], and data is scarce in equids. Here, our estimation of the rate of IgG antibodies loss (seroreversion parameter) suggests a lifelong carriage of such antibodies in many individuals, even though seroreversions remain possible. Therefore, these individuals may benefit from a long-term protection against WNV symptoms and contribute to herd immunity.

The estimation of the relative risk of infection in chickens as compared to horses (β) was not conclusive, probably because, in our dataset, the time overlap between sampling periods in horses and in chickens was short and with a low-level WNV circulation. Sampling longitudinally both species during an outbreak or as part of an infection experiment would allow assessing this parameter by comparing FOI applying to both species simultaneously. It would be an important metric to consider when comparing different sentinel surveillance strategies, and to prioritize either horses or domestic birds surveillance [85]. At equal *Culex* vector densities in the environment, hosts’ relative risk of infection notably depends on mosquitoes feeding preferences. While *Cx. quinquefasciatus* and *Cx. nigripalpus* are the two main candidate vectors for WNV in Guadeloupe Island [80], the former has been suggested to bite birds (including chickens) more than mammals, whereas it might be the opposite for the latter [73,86–89]. However, host feeding preferences have been shown to depend on environmental factors (urban vs. rural settings, hygrometry, etc.) and host availability [73], and they have yet to be fully determined for *Culex* spp. in Guadeloupe. *Cx. quinquefasciatus* is a vector commonly found in domestic areas close to human habitations, where it breeds in artificial containers, while *Cx. nigripalpus* is more often associated with peridomestic environments, rural and natural biotopes. Feeding preferences may also change with host skin surface area availability – which depends on animals’ size – rather than at the host individual level [90].

Our study quantified the regular circulation of WNV in Guadeloupe after 2002, based on sentinel surveillance data, despite no clinical report in humans or horses before 2024. However, it did not allow to determine whether it was due to series of virus introductions or as a result of a local enzootic circulation. Although no blood samples were collected earlier than July 2002, an earlier circulation of WNV on the island cannot be ruled out since a West Nile human case was detected as early as August 2001 in the Cayman Islands, a northern Caribbean territory [91]. From our median estimates of WNV FOI (e.g. in 2007 and 2012), the annual incidence rate in horses during outbreak years can be estimated to 5%-7%. Considering a total estimated equine population between ~500 (in 2003-2004) and ~1,000 (in 2017) on the island [80,92] – which is an overestimation of the susceptible equine population – and a proportion of neurological symptoms of ~10% of WNV infected horses [93,94], we could expect up to ~3-7 equine neurological disease cases in 2007 and 2012. Therefore, the lack of WNV clinical case evidence in both domestic animals and humans until 2024 may suggest a low sensitivity of WNV passive surveillance [38]. Indeed, in both horses and humans in the Caribbean, it may be jeopardized by the frequent co-occurrence of other pathogens with similar pathogenesis (e.g. equine piroplasmosis) and serological cross-reactivity for flaviviruses such as dengue [95,96]. This highlights the potential of a complementary sentinel WNV serological surveillance scheme in domestic animals, subject to the results of a more thorough costs-benefits analysis, as well as the importance of cross-sectoral collaborations.

In the future, building a mechanistic model fitted to infection data in vectors and wild birds would allow to unravel the processes underlying temporal changes in WNV FOI [97]. Based on such a model, a simulation study would help to determine what cost-efficient surveillance strategies (involving a seasonal component or not) could be implemented in the Caribbean to monitor WNV emergence or re-emergence [85], and therefore to mitigate its impacts on human and animal health.

## Acknowledgements

This work was supported by the BCOMING project (Horizon Europe project 101059483) funded by the European Union, by a DIM1Health postdoctoral fellowship awarded by the Conseil Régional d’Ile-de-France and by the EU project MALIN and the Guadeloupe Regional Council under the European Research and Development Funds (ERDF) 2014-2020 program (Grant 2018-FED-1084). Surveillance data collection was funded by local veterinary services (DAAF971). The authors would like to thank Mariana Geffroy, for her contribution in the organization of the WNV sero-surveillance data, as well as Thierry Lefrançois, Nathalie Vachiéry and Emmanuel Albina (CIRAD, Astre) for their contribution in establishing and/or reinforcing WNV active surveillance schemes in Guadeloupe.

## Data availability statement

Data is presented within the paper and its supplementary material. Code reproducing the article is available from the following link: https://github.com/JonathanBas/West_Nile_model_Guadeloupe.

## Author contributions

Conceptualization: VC, SL, JB, BD. Investigation (data collection): JP, NP, SL. Data Curation: JP, NP, SL. Funding Acquisition: VC, SL, BD. Formal Analysis: CH, JB. Software: CH, JB. Supervision: SL, JB, BD. Writing – Original Draft Preparation: JB. Writing – Review × Editing: All authors.

