## Supplementary Information for "Reconstructing the silent circulation of West Nile Virus in a Caribbean island during 15 years using sentinel serological data"

**Supplementary Table S1.** Formula of the force of infection depending on the serological model scenario and the species.

| Model scenario | Species | Force of infection λ(t) |
|---|---|---|
| **FlatStable** | Horses | $\Lambda$ |
| | Chickens | $\beta.\Lambda$ |
| **FlatVary** | Horses | $\Lambda(y(t))$ |
| | Chickens | $\beta.\Lambda(y(t))$ |
| **SeasoStable** | Horses | $\frac{\Lambda}{2}(1-\varepsilon)\left(1+cos\left(\frac{2\pi}{52}\ (t-\delta)\right)\right)+\varepsilon.\Lambda$ |
| | Chickens | $\beta\left[\frac{\Lambda}{2}(1-\varepsilon)\left(1+cos\left(\frac{2\pi}{52}\ (t-\delta)\right)\right)+\varepsilon.\Lambda\right]$ |
| **SeasoVary** | Horses | $\frac{\Lambda(y(t))}{2}(1-\varepsilon)\left(1+cos\left(\frac{2\pi}{52}\ (t-\delta)\right)\right)+\varepsilon.\Lambda(y(t))$ |
| | Chickens | $\beta\left[\frac{\Lambda(y(t))}{2}(1-\varepsilon)\left(1+cos\left(\frac{2\pi}{52}\ (t-\delta)\right)\right)+\varepsilon.\Lambda(y(t))\right]$ |

**Supplementary Note 1.**

To initialize the model and infer the true serological status of the first sample in each individual $i$, we introduced a parameter $NPV_1$, the negative predictive value of the first sample result for any individual (main manuscript, Table 1). By definition [1]:

$$NPV_1 = \frac{1 - P_1}{1 - P_1 + (1 - \eta).P_1} \qquad (S1)$$

Where $P_1$ was a parameter varying between 0 and 1 (it was the seroprevalence when individuals were first sampled). It was assumed to follow a Beta distribution of estimated parameters $\alpha_1$ and $\alpha_2$:

$$P_1 \sim Beta(\alpha_1, \alpha_2) \qquad (S2)$$

**Supplementary Note 2.**

Step 1 of the model fitting procedure consisted to fit a model analogous to the "SeasoStable" model to the mosquito abundance data, in order to obtain estimates of parameters $\delta$ and $\varepsilon$, which were used in turn as priors for Step 2. The abundance of mosquitoes trapped at a site j on a week t was modelled as:

$$N_{mosq,j}(t) \sim Normal\left(E_{mosq,j}(t), \sigma_{mosq}\right)$$

Where:

$$E_{mosq,j}(t) = N_0 F_j \left( (1 - \varepsilon)\frac{1}{2}\left(1 + cos\left(\frac{2\pi}{52}\,(t - \delta)\right)\right) + \varepsilon \right)$$

Where $N_0$ was a scale parameter, $F_j$ was the fixed effect of site $j$ (with Site 1 as reference, i.e. $F_1 = 1$), and $\delta$ and $\varepsilon$ were as defined in the main manuscript. All parameters were estimated with mostly uninformative priors (Supplementary Table S2).

**Supplementary Table S2.** Prior distributions used in Steps 1 and 2 of model fitting. Posterior distributions estimated for $\varepsilon$ and $\delta$ following Step 1 (i.e. the fit of the seasonal model to mosquito abundance data) were then used as informative prior distributions for Step 2 (i.e. the fit of all models to serological data).

| Parameter | Prior in Step 1<br>Fit to mosquito abundance data | Prior in Step 2<br>Fit to serological data |
|---|---|---|
| $\beta$ | - | Uniform(0, 10) |
| $\Lambda(y(t))$ or $\Lambda$ | - | Exponential transformation of Uniform(-20, 3) |
| $\varepsilon$ | Uniform(0, 1) | Beta(3.80, 22.43) |
| $\delta$ | Uniform(1, 53) | Normal(45.26, 0.988) |
| $\mu$ | - | Uniform(0, 0.5) |
| $\eta$ | - | Uniform(0, 1) |
| $NPV_1$ | - | Depends on $\eta$, $\alpha_1$ and $\alpha_2$ (see Supplementary Note 1) |
| $\alpha_1$ | - | Uniform(0, 100) |
| $\alpha_2$ | - | Uniform(0, 100) |
| $\sigma_{mosq}$ | Uniform(0, 100) | - |
| $N_0$ | Exponential transformation of Uniform(-5, 10) | - |
| $F_j$ | Uniform(0, 100) | - |

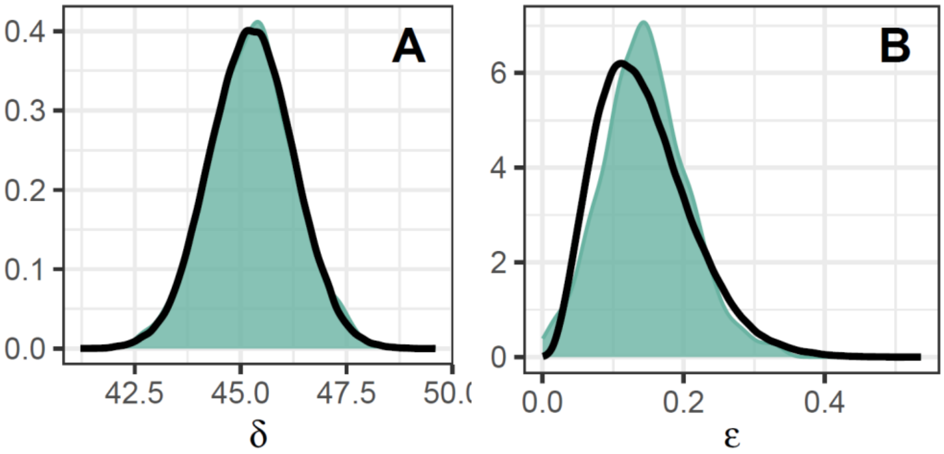


**Supplementary Figure S1.** Panel A: Posterior distribution of *δ* following Step 1 (colored area), and corresponding Normal distribution (of mean 45.26 and standard deviation 0.988) used as prior distribution in Step 2 (black line). Panel B: Posterior distribution of *ε* following Step 1 (colored area), and corresponding Beta distribution (of parameters 3.80 and 22.43) used as prior distribution in Step 2 (black line).

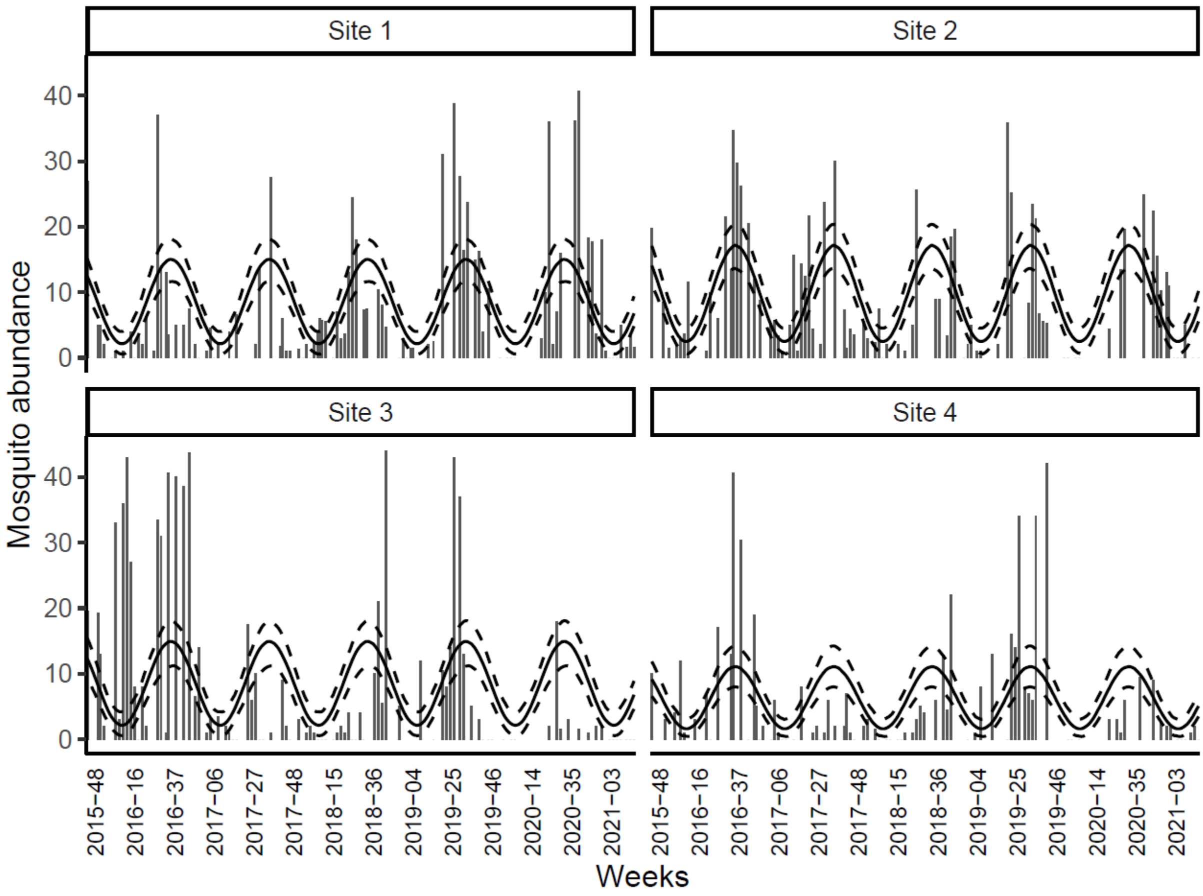


**Supplementary Figure S2.** Mosquito abundance in four collection sites in Guadeloupe Island between November 2015 and May 2021: observed data (vertical bars) and model predictions following Step 1 (median of 500 model repetitions: solid line; 95% prediction interval: dashed lines).

**Supplementary Table S3.** Values of the Deviance Information Criterion (DIC) for the different serological model scenarios.

| **Model scenario** | FlatStable | FlatVary | SeasoStable | SeasoVary |
|---|---|---|---|---|
| **DIC** | 1085 | 936 | 1072 | 926 |

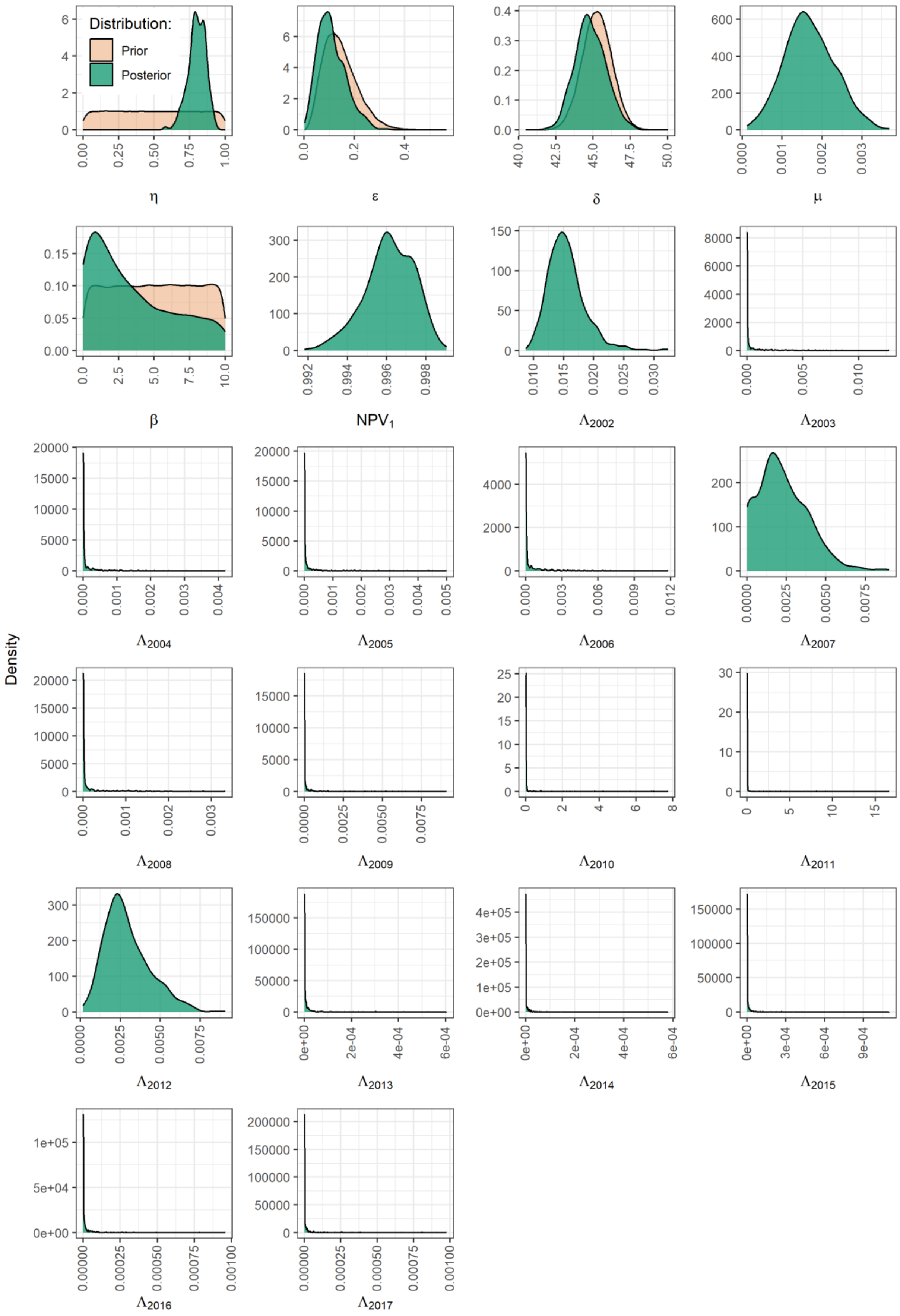
Distribution:
Prior
Posterior
Density
η
ε
δ
μ
β
NPV1
Λ2002
Λ2003
Λ2004
Λ2005
Λ2006
Λ2007
Λ2008
Λ2009
Λ2010
Λ2011
Λ2012
Λ2013
Λ2014
Λ2015
Λ2016
Λ2017

**Supplementary Figure S3.** Step 2 of the fitting process for the "SeasoVary" model (fit to the serological data): prior and posterior distributions of parameters. Here, the prior distribution is displayed only when it does not preclude the good visualization of the posterior distribution.
